# Partial and Radical cystectomy Provides Equivalent Oncologic Outcomes in T2 Bladder Cancer: a Population-Based Study

**DOI:** 10.1101/2022.05.07.22274793

**Authors:** Gongwei Long, Zhiquan Hu, Zheng Liu, Zhangqun Ye, Shaogang Wang, Dongwen Wang, Chunguang Yang

## Abstract

**Purpose:** To compare the oncologic outcomes of T2 bladder cancer (BCa) patients after partial cystectomy (PC) or radical cystectomy (RC).

**Materials and Methods:** Relevant data of T2 BCa patients diagnosed between 2004 and 2015 were retrieved from the Surveillance, Epidemiology, and End Results database. After propensity score matching, the oncologic outcomes after PC or RC were compared in patients who underwent lymph node dissection (LND) or not. Additionally, the benefits of different LND strategies were also explored.

**Results:** Eventually, 3834 T2 BCa cases were enrolled in the analysis, including 724 (18.9%) received PC cases and 3110 (81.1%) RC cases. The PC and RC groups presented entirely different profiles in clinical parameters such as age, number of lymph nodes (LNs) removed, and adjuvant therapy. Particularly, LNDs were performed in 92.0% of RC cases, while only in 45.4% of PC cases. After propensity score matching, PC and RC present similar oncologic outcomes regardless of the LND strategies. Further exploration found that LND could improve the prognosis of patients and the benefit is associated with the number of LNs removed.

**Conclusion:** PC and RC could provide equivalent oncologic outcomes in T2 BCa. LND is essential in curative surgery and could significantly affect the prognosis, but it was frequently neglected, especially in PC. The criteria for PC in T2 BCa need further exploration in future studies.

## INTRODUCTION

Bladder cancer (BCa) is one of the most common urological malignant diseases worldwide with high recurrence and progression rates (1). For non-muscle-invasive bladder tumors (NMIBC) contained in mucosa or submucosa, transurethral resection of bladder tumors (TURBT) is the standard treatment option (2, 3). But when tumors invaded the muscle layer, radical cystectomy (RC) combined with lymph node dissection (LND) is the standard surgical treatment (4, 5).

However, RC is the most morbid and complex urologic surgery. Over half of patients would experience postoperative complications and about 1-4% of patients might die within 90 days after RC (6, 7). Partial cystectomy (PC) is an alternative option for patients with a solitary and primary tumor without concomitant carcinoma in situ (CIS) that is amenable to resection with a 2 cm surgical margin, and about 5% to 10% of muscle-invasive BCa (MIBC) meet the criteria (8, 9). And it should be noted that the partial cystectomy rate decreased with time (10).

Cancer control is the main concern when performing the PC. Here we retrieved data from the Surveillance, Epidemiology, and End Results (SEER) database, and compare the oncologic outcomes after PC or RC to confirm the feasibility of PC in T2 BCa.

## METHODS

### Data source and cases inclusion

The data were retrieved from the SEER database, which is a population-based cancer database found by the National Cancer Institute that collects data on cancer incidence and survival data from US cancer registries.

We obtained BCa patients’ data diagnosed between 2004 and 2015 from 18 registries of the SEER database (November 2020 Submission) using the SEER*Stat 8.3.9 program. The process of screening patients was shown in Figure 1. The inclusion criteria were as follows: Urinary bladder cancer according to International Classification of Diseases for Oncology, 3rd edition (ICD-O3); Age ≥ 18 yr; diagnosed between 2004-2015; no distant metastasis; stage T2; underwent RC or PC; the information of LND is available.

**Figure 1:**
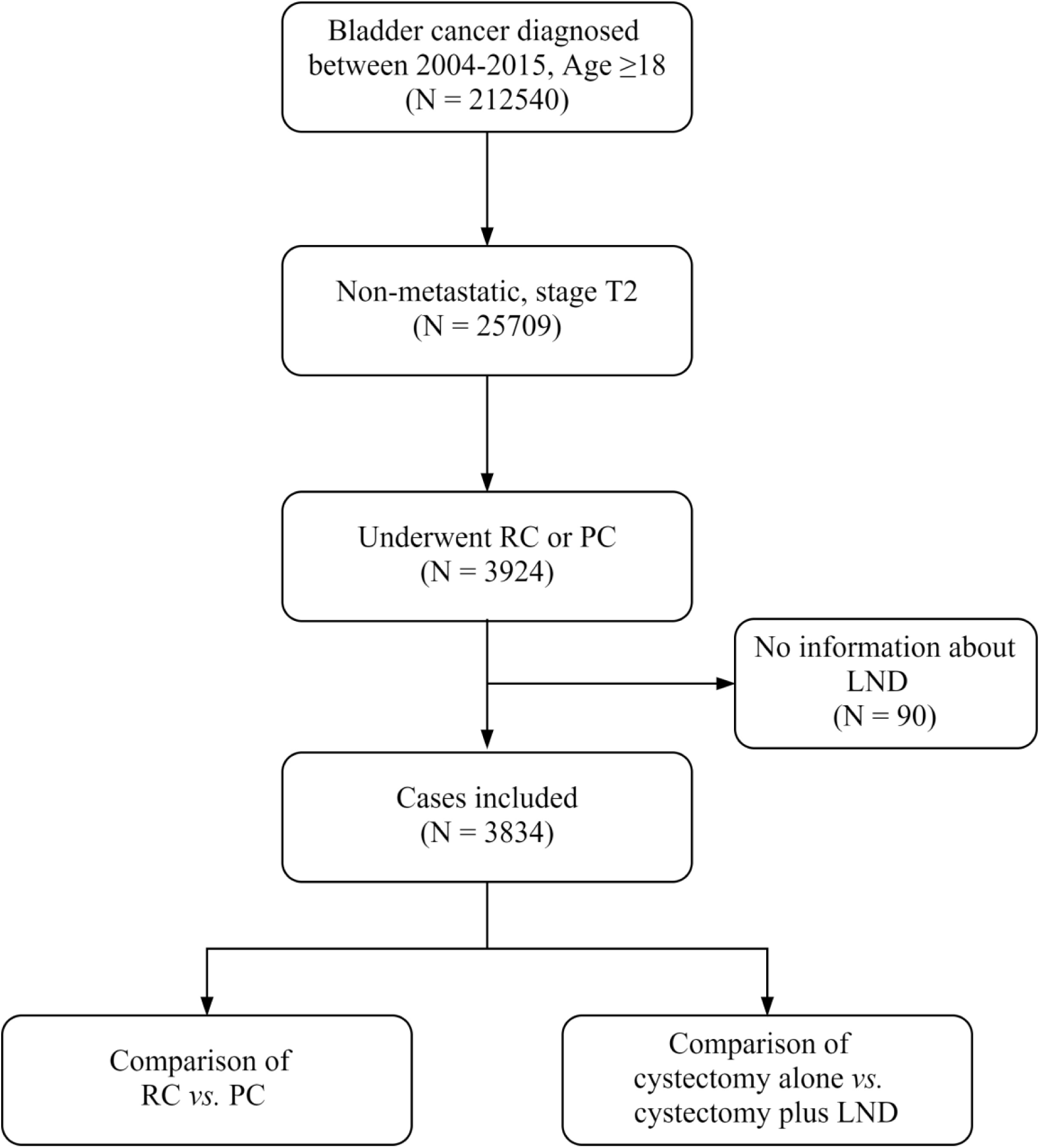
Flow chart of the patient screening. RC, radical cystectomy; PC, partial cystectomy; LND, lymph node dissection.

### Propensity score matching

Propensity score matching (PSM) is a method to reduce bias in observational studies where randomized treatment assignment is not possible. The R package “MatchIt” was used to perform nearest neighbor PSM with a ratio of 1:1(11).

To compare the oncologic outcomes after RC or PC, PSMs were performed separately in patients who underwent LND or not. Besides, to explore the effect of LND on oncologic outcomes, a PSM based on LND status was also performed.

### Statistical analysis

The statistical analysis was conducted using the R 4.0.2 software. Continuous data were presented as median and interquartile range (IQR) and compared using Student’s t or Wilcoxon rank-sum test. Categorical data were presented as count and percentage and compared with the Chi-square or Fisher exact test.

The overall survival (OS) and cancer-specific survival (CSS) were defined as the endpoints of oncologic outcomes. The Kaplan-Meier (K-M) curves were plotted and the log-rank tests were conducted to compare the survival of different groups with the Graphpad Prism 8.0.1 software. Univariate and multivariate Cox regression analyses were also performed to identify prognostic factors. Parameters with a P-value less than 0.05 in univariate analysis were included in multivariate analysis.

## RESULTS

### Characteristics of included patients

After screening, 3834 cases were enrolled in the analysis. The median (IQR) age of the included patients is 67 (60-76) yr. Among these patients, 724 (18.9%) patients received PC, while 3110 (81.1%) patients received RC. The baseline characteristics were summarized in Table 1.

**Table 1.**
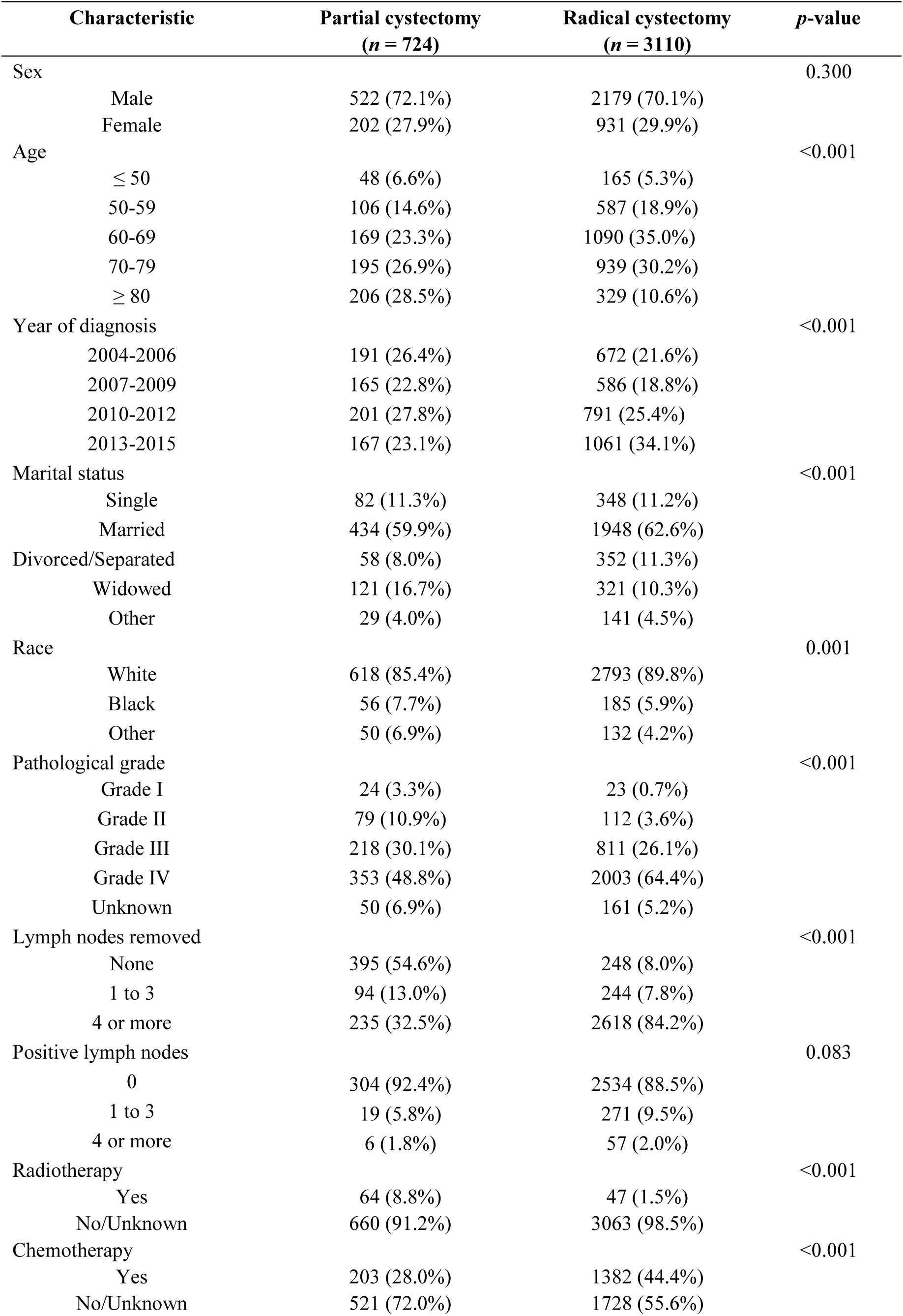
Characteristics of included T2 bladder cancer patients.

| Characteristic |  | Partial cystectomy<br>(*n* = 724) | Radical cystectomy<br>(*n* = 3110) | *p*-value |
| --- | --- | --- | --- | --- |
| Sex |  |  |  | 0.300 |
|  | Male | 522 (72.1%) | 2179 (70.1%) |  |
|  | Female | 202 (27.9%) | 931 (29.9%) |  |
| Age |  |  |  | <0.001 |
|  | ≤ 50 | 48 (6.6%) | 165 (5.3%) |  |
|  | 50-59 | 106 (14.6%) | 587 (18.9%) |  |
|  | 60-69 | 169 (23.3%) | 1090 (35.0%) |  |
|  | 70-79 | 195 (26.9%) | 939 (30.2%) |  |
|  | ≥ 80 | 206 (28.5%) | 329 (10.6%) |  |
| Year of diagnosis |  |  |  | <0.001 |
|  | 2004-2006 | 191 (26.4%) | 672 (21.6%) |  |
|  | 2007-2009 | 165 (22.8%) | 586 (18.8%) |  |
|  | 2010-2012 | 201 (27.8%) | 791 (25.4%) |  |
|  | 2013-2015 | 167 (23.1%) | 1061 (34.1%) |  |
| Marital status |  |  |  | <0.001 |
|  | Single | 82 (11.3%) | 348 (11.2%) |  |
|  | Married | 434 (59.9%) | 1948 (62.6%) |  |
|  | Divorced/Separated | 58 (8.0%) | 352 (11.3%) |  |
|  | Widowed | 121 (16.7%) | 321 (10.3%) |  |
|  | Other | 29 (4.0%) | 141 (4.5%) |  |
| Race |  |  |  | 0.001 |
|  | White | 618 (85.4%) | 2793 (89.8%) |  |
|  | Black | 56 (7.7%) | 185 (5.9%) |  |
|  | Other | 50 (6.9%) | 132 (4.2%) |  |
| Pathological grade |  |  |  | <0.001 |
|  | Grade I | 24 (3.3%) | 23 (0.7%) |  |
|  | Grade II | 79 (10.9%) | 112 (3.6%) |  |
|  | Grade III | 218 (30.1%) | 811 (26.1%) |  |
|  | Grade IV | 353 (48.8%) | 2003 (64.4%) |  |
|  | Unknown | 50 (6.9%) | 161 (5.2%) |  |
| Lymph nodes removed |  |  |  | <0.001 |
|  | None | 395 (54.6%) | 248 (8.0%) |  |
|  | 1 to 3 | 94 (13.0%) | 244 (7.8%) |  |
|  | 4 or more | 235 (32.5%) | 2618 (84.2%) |  |
| Positive lymph nodes |  |  |  | 0.083 |
|  | 0 | 304 (92.4%) | 2534 (88.5%) |  |
|  | 1 to 3 | 19 (5.8%) | 271 (9.5%) |  |
|  | 4 or more | 6 (1.8%) | 57 (2.0%) |  |
| Radiotherapy |  |  |  | <0.001 |
|  | Yes | 64 (8.8%) | 47 (1.5%) |  |
|  | No/Unknown | 660 (91.2%) | 3063 (98.5%) |  |
| Chemotherapy |  |  |  | <0.001 |
|  | Yes | 203 (28.0%) | 1382 (44.4%) |  |
|  | No/Unknown | 521 (72.0%) | 1728 (55.6%) |  |

As shown in Table 1, the PC and RC group had entirely different profiles in several clinical parameters including age (*p* < 0.001), year of diagnosis (*p* < 0.001), marital status (*p* < 0.001), race (*p* = 0.001), number of lymph nodes (LNs) removed (*p* < 0.001), radiotherapy (*p* < 0.001), and chemotherapy (*p* < 0.001).

Concretely speaking, the patients who underwent PC had a larger median age than those who underwent RC (72.0 *vs*. 67.0 yr, *p* < 0.001). The number of PC surgeries was relatively stable in periods between 2004 to 2015, while the number of RC had an increment. More married, separated, or divorced patients received RC, while more widowed patients underwent PC (*p* < 0.001). White patients received more RC, and black and minority patients preferred PC. The pathological grade IV was also more frequent in RC group (64.4% *vs*. 48.8%, *p* < 0.001).

LNDs were performed in 92.0% of patients who performed RC, while the rate in PC was only 45.4% (*p* < 0.001). Besides, the average number of LNs examined was higher in RC (16.1 *vs*. 4.2, *p* < 0.001). Subsequently, the LNs positive rate was also higher in RC group (11.5% *vs*. 7.6%, *p* = 0.034).

Additionally, adjuvant therapy profiles differed in groups. The PC group received more radiotherapy while the RC group underwent more chemotherapy.

In the cohort containing 3834 cases, RC was associated with a better OS (*p* < 0.001) and CSS (*p* = 0.047, Supplement Figure S1 A-B). However, considering the unbalanced baseline characteristics, a direct comparison of PC and RC could be severely biased due to these confounding factors. Subgroup analyses based on the LND were also performed, and interestingly, the OS and CSS of RC and PC were similar in different subgroups (Supplement Figure S1 C-F).

### Comparison of surgery strategies by PSM

In the 3834 included cases, 643 patients did not undergo LND and the other 3191 performed LND. After separate PSM, the baseline characteristics of PC and RC groups turned similar (Table S1).

In patients who underwent LND, PC plus LND had a median OS of 113.0 months and RC plus LND had a median OS of 92.0 months. The OS did not differ between the two surgery groups (Hazard Ratio (HR) 0.872, *p* = 0.212, Figure 2A). The CSS comparison also did not suggest a significant difference (HR 0.756, *p* = 0.072, Figure 2B).

**Figure 2:**
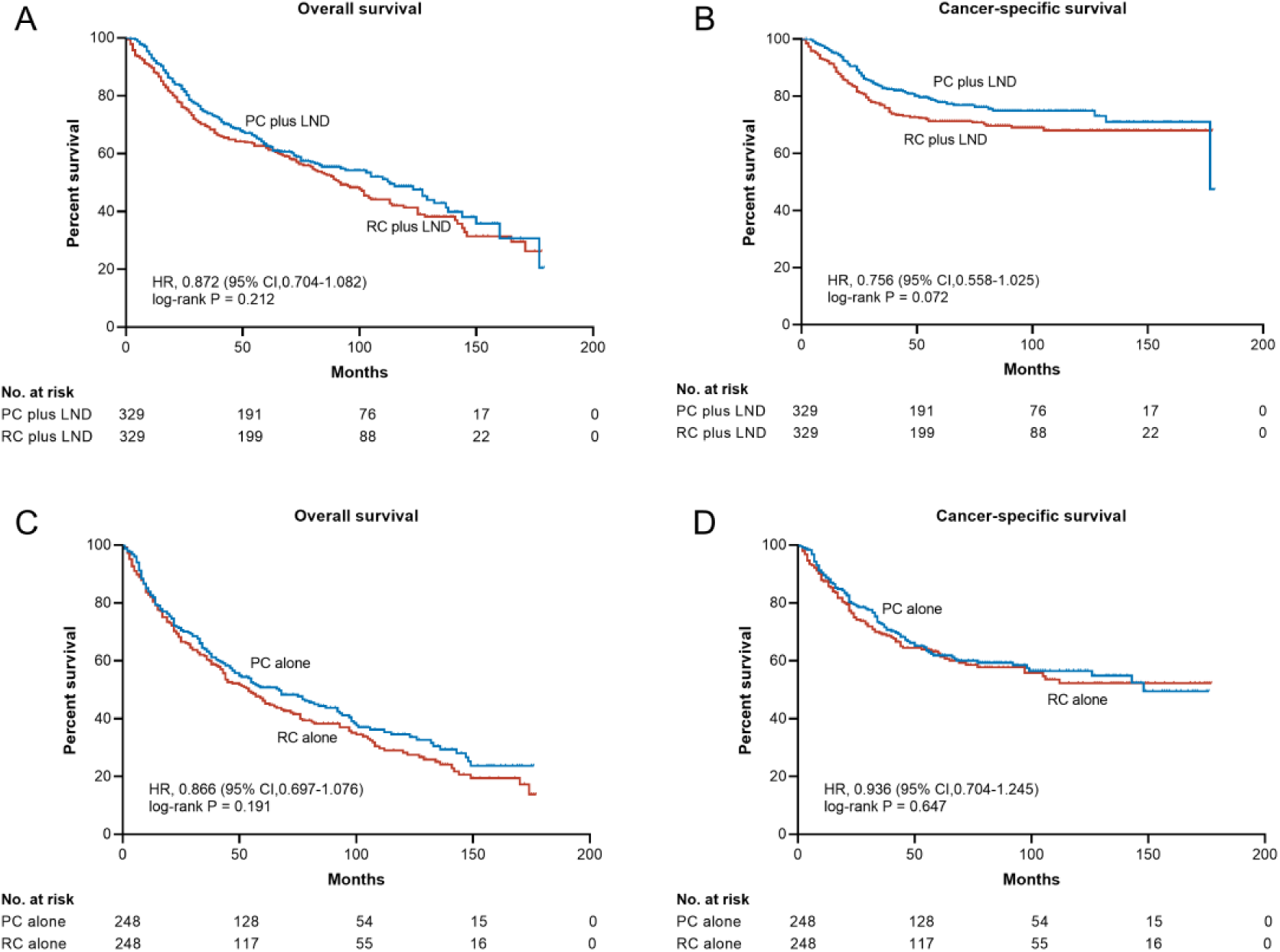
The KM plot showing oncologic outcomes after different surgeries. RC, radical cystectomy; PC, partial cystectomy; LND, lymph node dissection.

The median OS of patients who performed PC alone was 67.0 months, while that of patients who performed RC alone was 54.0 months. Similarly, the OS (HR 0.866, *p* = 0.191, Figure 2C) and CSS (HR 0.936, *p* = 0.647, Figure 2D) did not differ between the two surgery groups.

Notably, the OS was shorter in patients who did not perform LND (67.0 vs. 113.0 months in PC; 54.0 vs. 92.0 months in RC). To further explore the effect of LND on prognosis, another PSM was performed in the 3834 included cases. As seen in Supplement Table S2, patients were divided into three groups according to the number of LNs removed, and baseline characteristics were balanced after PSM. Either RC or PC was conducted in these patients, and the proportions of PC were around 30% and similar in the three groups. In patients who underwent LND, the LN positive rates were also balanced in groups with different numbers of LNs removed.

The KM plots in Figure 3 suggested that the removal of 1-3 LNs could already prolong the OS (HR 0.808, *p* = 0.027) and CSS (HR 0.755, *p* = 0.032) of T2 BCa patients. Besides, removing 4 or more LNs also suggested advantages in OS (HR 0.798, *p* = 0.026) over 1-3 LNs, but not statistically significant in CSS (HR 0.785, *p* = 0.091).

**Figure 3:**
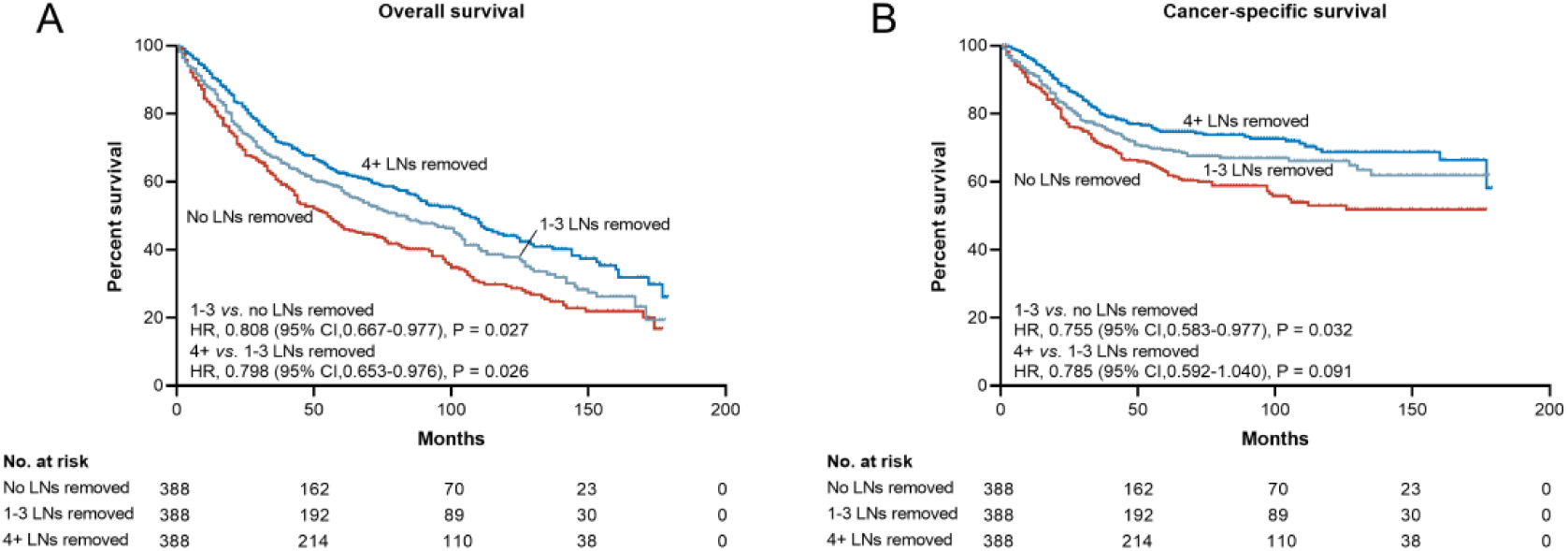
The KM plot showing oncologic outcomes of different lymph node dissection strategies. LN, lymph node.

### Prognostic factors after surgery

To further explore the prognostic factors after surgery in T2 patients, we also conducted Cox regression analysis in the 3834 included cases.

In the analysis of OS in patients who performed LND, the age, diagnosis year, marital status, race, number of LNs removed, number of positive LNs, radiotherapy, and chemotherapy were associated with the OS (Table 2). The death risk increased along with the age and decreased along with the year of diagnosis. Positive LNs are associated with shorter survival. Similar outcomes were also found in the CSS analysis (Supplement Table S3).

**Table 2.**
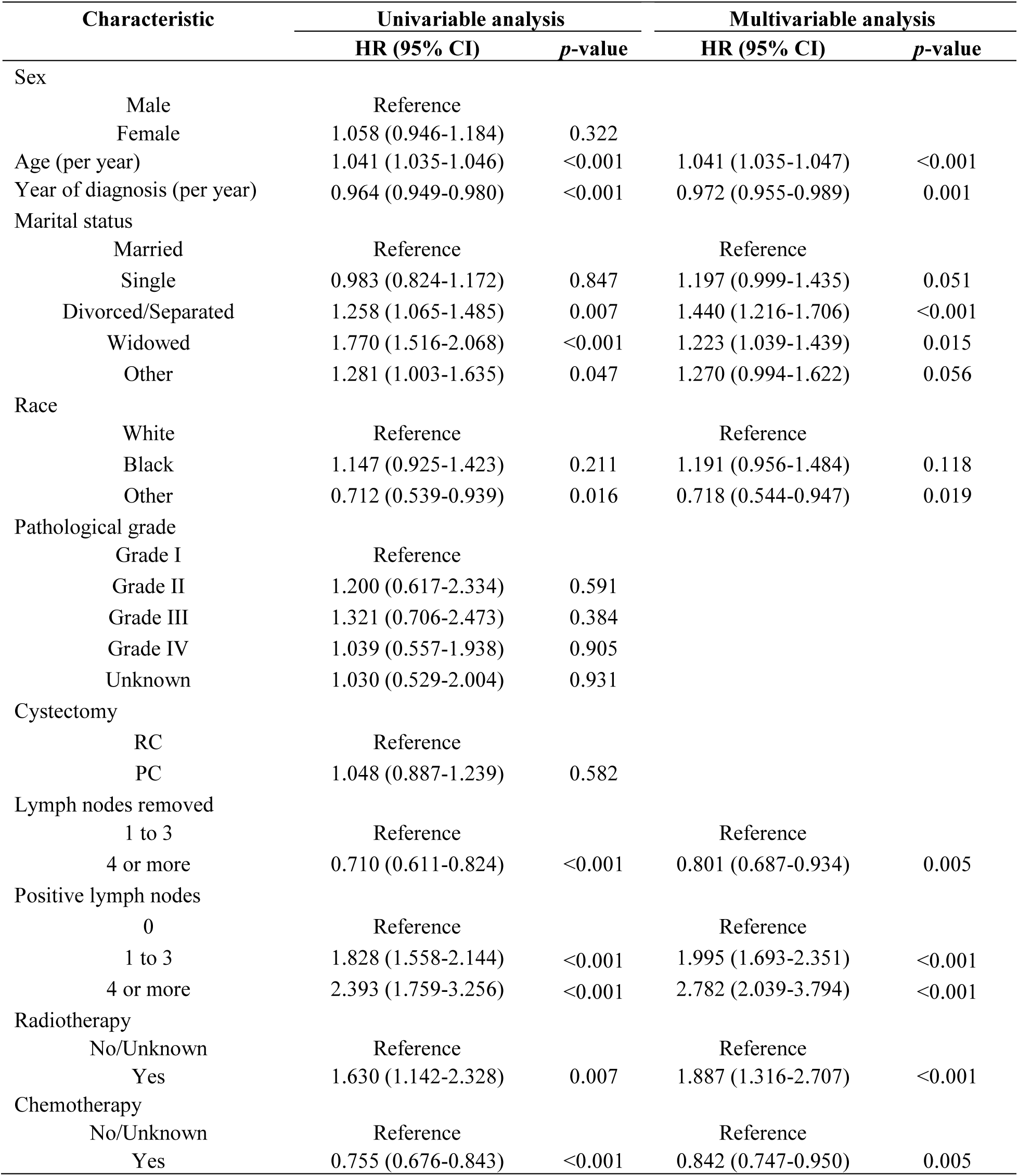
Univariate and multivariate Cox regression analysis of OS in patients who underwent LND.

| Characteristic | Univariable analysis |  | Multivariable analysis |  |
| --- | --- | --- | --- | --- |
|  | HR (95% CI) | *p*-value | HR (95% CI) | *p*-value |
| Sex |  |  |  |  |
| Male | Reference |  |  |  |
| Female | 1.058 (0.946-1.184) | 0.322 |  |  |
| Age (per year) | 1.041 (1.035-1.046) | <0.001 | 1.041 (1.035-1.047) | <0.001 |
| Year of diagnosis (per year) | 0.964 (0.949-0.980) | <0.001 | 0.972 (0.955-0.989) | 0.001 |
| Marital status |  |  |  |  |
| Married | Reference |  | Reference |  |
| Single | 0.983 (0.824-1.172) | 0.847 | 1.197 (0.999-1.435) | 0.051 |
| Divorced/Separated | 1.258 (1.065-1.485) | 0.007 | 1.440 (1.216-1.706) | <0.001 |
| Widowed | 1.770 (1.516-2.068) | <0.001 | 1.223 (1.039-1.439) | 0.015 |
| Other | 1.281 (1.003-1.635) | 0.047 | 1.270 (0.994-1.622) | 0.056 |
| Race |  |  |  |  |
| White | Reference |  | Reference |  |
| Black | 1.147 (0.925-1.423) | 0.211 | 1.191 (0.956-1.484) | 0.118 |
| Other | 0.712 (0.539-0.939) | 0.016 | 0.718 (0.544-0.947) | 0.019 |
| Pathological grade |  |  |  |  |
| Grade I | Reference |  |  |  |
| Grade II | 1.200 (0.617-2.334) | 0.591 |  |  |
| Grade III | 1.321 (0.706-2.473) | 0.384 |  |  |
| Grade IV | 1.039 (0.557-1.938) | 0.905 |  |  |
| Unknown | 1.030 (0.529-2.004) | 0.931 |  |  |
| Cystectomy |  |  |  |  |
| RC | Reference |  |  |  |
| PC | 1.048 (0.887-1.239) | 0.582 |  |  |
| Lymph nodes removed |  |  |  |  |
| 1 to 3 | Reference |  | Reference |  |
| 4 or more | 0.710 (0.611-0.824) | <0.001 | 0.801 (0.687-0.934) | 0.005 |
| Positive lymph nodes |  |  |  |  |
| 0 | Reference |  | Reference |  |
| 1 to 3 | 1.828 (1.558-2.144) | <0.001 | 1.995 (1.693-2.351) | <0.001 |
| 4 or more | 2.393 (1.759-3.256) | <0.001 | 2.782 (2.039-3.794) | <0.001 |
| Radiotherapy |  |  |  |  |
| No/Unknown | Reference |  | Reference |  |
| Yes | 1.630 (1.142-2.328) | 0.007 | 1.887 (1.316-2.707) | <0.001 |
| Chemotherapy |  |  |  |  |
| No/Unknown | Reference |  | Reference |  |
| Yes | 0.755 (0.676-0.843) | <0.001 | 0.842 (0.747-0.950) | 0.005 |

Notably, Radiotherapy was associated with increased death risk (HR (95% CI), 1.887 (1.316-2.707); *p* < 0.001) while chemotherapy reduced the risk (HR (95% CI), 0.842 (0.747-0.950); *p* = 0.005). In the CSS analysis (Supplement Table S3), the chemotherapy was no longer associated with a better prognosis, while the radiotherapy was still a risk factor for survival (HR (95% CI), 2.526 (1.736-3.675); *p* < 0.001).

In patients who performed PC or RC alone, only age and radiotherapy were predictive for OS (Table 3) or CSS (Supplement Table S4). Similarly, in patients who underwent LND, elder patients (HR, 1.039, *p* < 0.001 in OS; HR, 1.023, *p* < 0.001 in CSS) and radiotherapy (HR, 1.414, *p* = 0.024 in OS; HR, 1.552, *p* = 0.023 in CSS) were also associated with poor OS and CSS.

**Table 3.** Univariate and multivariate Cox regression analysis of OS in patients without LND.

| Characteristic |  | Univariable analysis |  | Multivariable analysis |  |
| --- | --- | --- | --- | --- | --- |
|  |  | HR (95% CI) | p-value | HR (95% CI) | p-value |
| Sex |  |  |  |  |  |
|  | Male | Reference |  |  |  |
|  | Female | 0.994 (0.810-1.220) | 0.953 |  |  |
| Age (per year) |  | 1.039 (1.030-1.048) | <0.001 | 1.039 (1.029-1.048) | <0.001 |
| Year of diagnosis (per year) |  | 0.994 (0.966-1.024) | 0.706 |  |  |
| Marital status |  |  |  |  |  |
|  | Married | Reference |  | Reference |  |
|  | Single | 0.907 (0.656-1.255) | 0.556 | 1.146 (0.825-1.592) | 0.416 |
|  | Divorced/Separated | 1.017 (0.720-1.438) | 0.923 | 1.153 (0.815-1.635) | 0.426 |
|  | Widowed | 1.480 (1.169-1.872) | 0.001 | 1.077 (0.843-1.375) | 0.555 |
|  | Other | 0.881 (0.481-1.613) | 0.682 | 0.953 (0.520-1.746) | 0.875 |
| Race |  |  |  |  |  |
|  | White | Reference |  |  |  |
|  | Black | 0.930 (0.666-1.299) | 0.670 |  |  |
|  | Other | 0.644 (0.401-1.034) | 0.068 |  |  |
| Pathological grade |  |  |  |  |  |
|  | Grade I | Reference |  |  |  |
|  | Grade II | 0.656 (0.355-1.214) | 0.180 |  |  |
|  | Grade III | 1.068 (0.637-1.790) | 0.802 |  |  |
|  | Grade IV | 0.899 (0.540-1.495) | 0.681 |  |  |
|  | Unknown | 0.795 (0.430-1.470) | 0.465 |  |  |
| Cystectomy |  |  |  |  |  |
|  | RC | Reference |  |  |  |
|  | PC | 0.941 (0.776-1.141) | 0.537 |  |  |
| Radiotherapy |  |  |  |  |  |
|  | No/Unknown | Reference |  | Reference |  |
|  | Yes | 1.585 (1.176-2.137) | 0.002 | 1.414 (1.047-1.911) | 0.024 |
| Chemotherapy |  |  |  |  |  |
|  | No/Unknown | Reference |  |  |  |
|  | Yes | 0.893 (0.711-1.121) | 0.329 |  |  |

## DISCUSSION

Currently, the spectra of BCa management quite differ in NMIBC and MIBC. For NMIBC, conservative surgeries are recommended, and a piece by piece TURBT has long been the standard surgical treatment (2, 3). The *En bloc* resection allows complete resection of neoplastic and adjacent normal tissue, and is getting more popular recently due to its advantages for pathological staging and cancer control (12-14). But for MIBC, the standard treatment swiftly turns to RC, after which the quality of life of patients was severely impaired.

Bladder sparing options including TURBT and PC have been developed during the last decade. However, studies showed that about a third of patients who underwent TURBT were under-staged(15). PC is an alternative procedure for MIBC and it offers advantages such as complete tumor excision, adequate bladder, and accurate staging of both initial tumor and LN metastatic status. Many single-institutional series suggested that PC has acceptable oncologic outcomes in appropriately selected patients, while the perioperative complication rate decreased to less than 10% (8, 16, 17).

The main concern of bladder sparing therapy is the prognosis. In our analysis, the real-world data suggested that PC was associated with a much worse prognosis (Supplement Figure S1). But we should notice that LND is a main confounding factor. In patients who performed PC, the LNDs were only conducted in less than half of the patients, and this might be the reason for the worse prognosis of patients who received PC. In fact, the guidelines emphasized that a standard LND up to the common iliac bifurcation is mandatory during surgery with curative intent (4, 5). The optimal extent of LND is currently under debate. A report pointed out that LN metastases outside the boundaries of standard LND are common (18). Therefore, an extended LND might be better than the standard LND (19), but the recent randomized-controlled trial failed to show the superiority (20). In the present analysis, the regions of LND were not documented, and according to the number of LNs removed, most LND should be performed with the standard method. Our analysis found that whether the LND was performed and the number of LNs removed could affect the prognosis of patients (Figure 3). These findings illustrated that the LND is an integral part of cystectomy and should not be skipped when feasible.

After adjustment of confounding factors, there was no significant difference in oncologic outcomes after PC or RC. This result supported that, for patients who meet the criteria for PC, the bladder-sparing PC is adequate for cancer control. Many comparative studies also explored the oncologic efficacy of PC. Knoedler et al. performed a 1:2 matched case-control study to compare PC and RC, and concluded that no significant differences existed between PC and RC with regard to 10-year postoperative metastasis-free survival, CSS, or OS(21). Herr et al.(15) and Sternberg et al.(22) found that in these MIBC patients who responded to neoadjuvant chemotherapy, PC could achieve oncologic outcomes similar to RC. Besides these single-institutional reports, Capitanio et al.(23) and Mistretta et al.(24) utilized the SEER data in different periods and suggested that PC does not undermine cancer control. These two studies supported the feasibility of PC in non-metastatic MIBC, but it should be noticed that adjuvant therapy strategies were not balanced. Our analysis focused on the T2 patients and adjusted the confounding factors including radiotherapy and chemotherapy and the conclusion might be less biased.

Importantly, not all MIBC patients are suitable for the PC procedure, and the selection of patients is critical for oncologic outcomes. In the early 2000s, Memorial Sloan Kettering Cancer Center (MSKCC) and MD Anderson developed the PC criteria, and the indication was restricted to solitary tumors without CIS that allows resection with a 2 cm surgical margin, and MD Anderson emphasized the restriction for patients without the need for ureteral re-implantation in a normally functioning bladder (8, 16). Under such criteria, PC were surgically feasible for about 5% to 10% of MIBC(9). While in our analysis, the proportion of PC is near 20%. We only included the T2 BCa patients, which might be a reason. On the other hand, the criteria might not be strictly applied in clinical practice. But still, the PC showed non-inferior cancer control, implying that the criteria for PC in T2 patients could be more flexible.

Interestingly, in the Cox regression analysis, the effect of adjuvant therapies was controversial. In patients who underwent LND, chemotherapy was associated with improved OS, while radiotherapy was significantly associated with worse OS and CSS. Similar results were also reported by other analyses of the SEER database (25, 26). One possible explanation is that radiotherapy would be applied as a palliative treatment for patients who progressed. Currently, perioperative radiotherapy, especially preoperative, is not recommended for operable MIBC and its role needs further exploration.

This study was based on the national population data and could provide a valid interpretation for real-world practice. This study utilized the PSM method to conclude the clinical efficacy of different cystectomy surgeries and LND strategies. Otherwise, as a retrospective study, several limitations of this study should be noted such as selection bias and missing information. For example, patients who underwent PC are more likely to have solitary primary tumors without concomitant CIS, and tumor history, multifocality, and CIS have been proven as important prognostic factors (8, 16). Although the number of LNs removed was documented in the SEER database, the region of LND was not clear, thus preventing further exploration. Further randomized-controlled trials in patients suitable for PC are warranted to ascertain the cancer control of PC.

## CONCLUSION

In conclusion, our results suggested that PC and RC could provide equivalent oncologic outcomes in T2 BCa. On the other hand, LND could significantly affect the prognosis of patients and should be a mandatory part of cystectomy when surgical feasible.

## Data Availability

All data produced in the present study are available upon reasonable request to the authors

## REFERENCE

1. Bray F, Ferlay J, Soerjomataram I, Siegel RL, Torre LA and Jemal A. Global cancer statistics 2018: GLOBOCAN estimates of incidence and mortality worldwide for 36 cancers in 185 countries. CA Cancer J Clin. (2018) 68:394–424 doi:10.3322/caac.21492

2. Babjuk M, Burger M, Compérat EM, Gontero P, Mostafid AH, Palou J, et al. European Association of Urology Guidelines on Non-muscle-invasive Bladder Cancer (TaT1 and Carcinoma In Situ) - 2019 Update. Eur Urol. (2019) 76:639–657 doi:10.1016/j.eururo.2019.08.016

3. Chang SS, Boorjian SA, Chou R, Clark PE, Daneshmand S, Konety BR, et al. Diagnosis and Treatment of Non-Muscle Invasive Bladder Cancer: AUA/SUO Guideline. J Urol. (2016) 196:1021–9 doi:10.1016/j.juro.2016.06.049

4. Witjes JA, Bruins HM, Cathomas R, Compérat EM, Cowan NC, Gakis G, et al. European Association of Urology Guidelines on Muscle-invasive and Metastatic Bladder Cancer: Summary of the 2020 Guidelines. Eur Urol. (2021) 79:82–104 doi:10.1016/j.eururo.2020.03.055

5. Chang SS, Bochner BH, Chou R, Dreicer R, Kamat AM, Lerner SP, et al. Treatment of Non-Metastatic Muscle-Invasive Bladder Cancer: AUA/ASCO/ASTRO/SUO Guideline. J Urol. (2017) 198:552–559 doi:10.1016/j.juro.2017.04.086

6. Shabsigh A, Korets R, Vora KC, Brooks CM, Cronin AM, Savage C, et al. Defining early morbidity of radical cystectomy for patients with bladder cancer using a standardized reporting methodology. Eur Urol. (2009) 55:164–74 doi:10.1016/j.eururo.2008.07.031

7. Djaladat H, Katebian B, Bazargani ST, Miranda G, Cai J, Schuckman AK, et al. 90-Day complication rate in patients undergoing radical cystectomy with enhanced recovery protocol: a prospective cohort study. World J Urol. (2017) 35:907–911 doi:10.1007/s00345-016-1950-z

8. Kassouf W, Swanson D, Kamat AM, Leibovici D, Siefker-Radtke A, Munsell MF, et al. Partial cystectomy for muscle invasive urothelial carcinoma of the bladder: a contemporary review of the M. D. Anderson Cancer Center experience. J Urol. (2006) 175:2058–62 doi:10.1016/s0022-5347(06)00322-3

9. Peak TC and Hemal A. Partial cystectomy for muscle-invasive bladder cancer: a review of the literature. Transl Androl Urol. (2020) 9:2938–2945 doi:10.21037/tau.2020.03.04

10. Fedeli U, Fedewa SA and Ward EM. Treatment of muscle invasive bladder cancer: evidence from the National Cancer Database, 2003 to 2007. J Urol. (2011) 185:72–8 doi:10.1016/j.juro.2010.09.015

11. Ho D, Imai K, King G and Stuart EA. MatchIt: Nonparametric Preprocessing for Parametric Causal Inference. Journal of Statistical Software. (2011) 42:1 –28 doi:10.18637/jss.v042.i08

12. Teoh JY-C, MacLennan S, Chan VW-S, Miki J, Lee H-Y, Chiong E, et al. An International Collaborative Consensus Statement on En Bloc Resection of Bladder Tumour Incorporating Two Systematic Reviews, a Two-round Delphi Survey, and a Consensus Meeting. European urology. (2020) 78:546–569 doi:10.1016/j.eururo.2020.04.059

13. Long G, Zhang Y, Sun G, Ouyang W, Liu Z and Li H. Safety and efficacy of thulium laser resection of bladder tumors versus transurethral resection of bladder tumors: a systematic review and meta-analysis. Lasers Med Sci. (2021) 36:1807–1816 doi:10.1007/s10103-021-03272-7

14. Liu Z, Long G, Zhang Y, Sun G, Ouyang W, Wang S, et al. Thulium Laser Resection of Bladder Tumors vs. Conventional Transurethral Resection of Bladder Tumors for Intermediate and High Risk Non-Muscle-Invasive Bladder Cancer Followed by Intravesical BCG Immunotherapy. Front Surg. (2021) 8:759487 doi:10.3389/fsurg.2021.759487

15. Herr HW, Bajorin DF and Scher HI. Neoadjuvant chemotherapy and bladder-sparing surgery for invasive bladder cancer: ten-year outcome. J Clin Oncol. (1998) 16:1298–301 doi:10.1200/jco.1998.16.4.1298

16. Holzbeierlein JM, Lopez-Corona E, Bochner BH, Herr HW, Donat SM, Russo P, et al. Partial cystectomy: a contemporary review of the Memorial Sloan-Kettering Cancer Center experience and recommendations for patient selection. J Urol. (2004) 172:878–81 doi:10.1097/01.ju.0000135530.59860.7d

17. Smaldone MC, Jacobs BL, Smaldone AM and Hrebinko RL, Jr. Long-term results of selective partial cystectomy for invasive urothelial bladder carcinoma. Urology. (2008) 72:613–6 doi:10.1016/j.urology.2008.04.052

18. Dorin RP, Daneshmand S, Eisenberg MS, Chandrasoma S, Cai J, Miranda G, et al. Lymph node dissection technique is more important than lymph node count in identifying nodal metastases in radical cystectomy patients: a comparative mapping study. Eur Urol. (2011) 60:946–52 doi:10.1016/j.eururo.2011.07.012

19. Bruins HM, Veskimae E, Hernandez V, Imamura M, Neuberger MM, Dahm P, et al. The impact of the extent of lymphadenectomy on oncologic outcomes in patients undergoing radical cystectomy for bladder cancer: a systematic review. Eur Urol. (2014) 66:1065–77 doi:10.1016/j.eururo.2014.05.031

20. Gschwend JE, Heck MM, Lehmann J, Rübben H, Albers P, Wolff JM, et al. Extended Versus Limited Lymph Node Dissection in Bladder Cancer Patients Undergoing Radical Cystectomy: Survival Results from a Prospective, Randomized Trial. Eur Urol. (2019) 75:604–611 doi:10.1016/j.eururo.2018.09.047

21. Knoedler JJ, Boorjian SA, Kim SP, Weight CJ, Thapa P, Tarrell RF, et al. Does partial cystectomy compromise oncologic outcomes for patients with bladder cancer compared to radical cystectomy? A matched case-control analysis. J Urol. (2012) 188:1115–9 doi:10.1016/j.juro.2012.06.029

22. Sternberg CN, Pansadoro V, Calabrò F, Schnetzer S, Giannarelli D, Emiliozzi P, et al. Can patient selection for bladder preservation be based on response to chemotherapy? Cancer. (2003) 97:1644–52 doi:10.1002/cncr.11232

23. Capitanio U, Isbarn H, Shariat SF, Jeldres C, Zini L, Saad F, et al. Partial cystectomy does not undermine cancer control in appropriately selected patients with urothelial carcinoma of the bladder: a population-based matched analysist. Urology. (2009) 74:858–64 doi:10.1016/j.urology.2009.03.052

24. Mistretta FA, Cyr SJ, Luzzago S, Mazzone E, Knipper S, Palumbo C, et al. Partial Cystectomy With Pelvic Lymph Node Dissection for Patients With Nonmetastatic Stage pT2-T3 Urothelial Carcinoma of Urinary Bladder: Temporal Trends and Survival Outcomes. Clin Genitourin Cancer. (2020) 18:129-137.e3 doi:10.1016/j.clgc.2019.09.008

25. Wang W, Liu J and Liu L. Development and Validation of a Prognostic Model for Predicting Overall Survival in Patients With Bladder Cancer: A SEER-Based Study. Front Oncol. (2021) 11:692728 doi:10.3389/fonc.2021.692728

26. Zhan X, Guo J, Chen L, Deng W, Liu X, Zhu K, et al. Prognostic significance of bladder neck involvement in non-muscle-invasive bladder cancer: A SEER database analysis with 19,919 patients. Cancer Med. (2021) 10:6868–6880 doi:10.1002/cam4.4219

